# Child poverty and under-5 children services spending on early childhood development in England – a longitudinal local area ecological study of children born between 2017 and 2022

**DOI:** 10.1101/2025.09.05.25335147

**Authors:** Yu Wei Chua, Caitlin Murray, Davara Bennett, Gianmaria Niccodemi, Lateef Akanni, Luke Munford, Dougal Hargreaves, David Taylor-Robinson

**Author notes:** **Corresponding author:** Yu Wei Chua. **Competing interests:** We declare no competing interests. **Author contributions:** YWC is lead author and guarantor. Conceptualisation: YWC, DB, DTR; Methodology: YWC, DB, GN, LA, DTR; Formal analysis: YWC; Data Curation: YWC, CM; Writing – Original Draft: YWC, CM; Writing – Review & Editing: DB, GN, LA, DH, DTR, LM; Supervision: YWC; Funding acquisition: YWC, DTR. **Patient and public involvement:** No patient and public involvement activity was carried out for this study. **Data availability:** All data used in this study are publicly available.

## Abstract

**Background:** School readiness has fallen in the UK recently. We investigated the contribution of changes in child poverty and early years spending to trends in children not developmentally on track at 2-2.5 years.

**Methods:** In this longitudinal, ecological study of 143 local authorities (LA) in England, our outcomes were the rates per 100 children not on track in Any Domain, and in specific domains, assessed on the Ages and Stages Questionnaire 3 (ASQ-3), for children born between 2016/17 to 2021/22. Using within-between models controlling for confounders, we estimated associations between relative child poverty rates (<60% OECD median), under-5 children services spending, and free school meal eligibility (percentage eligibility in 5-to 11-year-old state-funded primary school pupils) in sensitivity analyses, on the outcomes.

**Findings:** Independent of spending, a 10 percentage-point (pp) increase in child poverty between LAs was associated with an increase in children not on track in Any Domain (9%[95%CI:1;18]), and in all domains except Gross Motor, but non-significant within-LA associations. Between-LA associations of free school meal eligibility were smaller and less precise. Within-LA, a 10pp free school meal eligibility increase was associated with a 16%[0;36] increase in Any Domain. Independent of child poverty, higher between-LA or within-LA spending of £100 per child under 5 were associated with around 4%[0; 8] relative decrease in Any Domain, and most strongly associated with Fine Motor. There was a protective effect of spending on reducing the size of the between-LA association between child poverty and rates of children developmentally on track.

**Interpretation:** Higher spending on early-years children’s services and lower child poverty were associated with better outcomes before school entry, and are key policy priorities to meet school readiness targets.

**Funding:** National institute for Health and Care Research (NIHR), NIHR Oxford Health Biomedical Research Consortium, NIHR School for Public Health Research, NIHR Applied Research Collaboration Greater Manchester

---

Early childhood development (ECD) sets the foundation for later learning and development, with long-lasting effects on adult health, behaviour, and economic productivity.^1,2^ One of the UK government’s current policy priorities is to remedy the poor state of children’s development by school entry age to “break down the barriers to opportunity”. Just over two-thirds of children in England are considered school ready by teachers,^3^ and the government aims to increase this to 75%. In addition, the UK has some of the best available data globally on the state of ECD before and after the pandemic.^4^ These recent trends show that ECD at 2 to 2.5 years was worse during and after the pandemic in Scotland and England,^4,5^ emphasising the need to prioritise relevant areas of investment in the post-pandemic political and economic climate to improve children’s development.

Child poverty and early interventions are two promising policy targets for improving child development. Although there is extensive evidence that child poverty harms early childhood developmental outcomes, including motor, socioemotional and cognitive domains^1,2,6^, there is limited evidence in the UK context on the impact of trends in child poverty on children’s early development (see Box 1). In the UK, most recent 2023/24 estimates indicate that around 30% of children live in relative poverty after housing costs.^7^ Child poverty was rising before the COVID-19 pandemic, against a backdrop of increased employment rates offset by a fall in benefits entitlement.^8^ During the pandemic, child poverty fell slightly due to the temporary universal credit uplift, and then returned to pre-pandemic levels.^9^ Child poverty is projected to rise further.^10^

Early interventions are effective for improving ECD,^11–13^ and often do so by addressing the needs of disadvantaged groups. National ECD programmes have been developed in several countries, including Sure Start in the UK, providing integrated services (e.g., early education, family and child health, and nutrition). Initial evaluations of Sure Start showed little evidence of benefits, but recent evidence important developmental outcomes particularly for disadvantaged groups. In the UK, there has also been substantial changes in funding for children’s services (see Appendix S1). Following drastic funding cuts to Sure Start children services since 2010/11,^14^ there were small increases in early intervention funding for under-5 children services during and after the pandemic.^15^ Given the role that children services play in delivering early interventions, these cuts are expected to harm children’s development. However, there is no suitable evidence to quantify, in a timely manner, the impact of cuts to children services in the UK (Box 1).

Up-to-date, timely, national evidence in the UK on how changes in poverty and spending are affecting ECD is lacking, but is crucial to inform policy decisions. Regional variation in child poverty trends is substantial,^16^ as is variation in how children services spending were managed by different local authorities.^14^ This study leverage recently available national-level data on ECD at 2 to 2.5 years and contextual differences (over time and between local authorities) in England. We investigate as a natural policy experiment at a local authority level, associations between child poverty and under-5 children services spending on ECD amongst children born before and during the COVID-19 pandemic.

## Box 1

**Research in context**

**Evidence before this study**

*Child poverty and ECD*

We examined the state of evidence and current research on the association between poverty and early childhood developmental outcomes through a literature search combining search terms for “early childhood”, “poverty” and “early childhood developmental outcomes”. We included reviews and primary studies (including study protocols) within the last 10 years (up to 16 August 2024) published in English. We searched Medline, CINAHL, Web of Science and PsychInfo databases. We examined evidence that defined poverty as a lack of material resources to meet need, excluding evidence examining socioeconomic status or disadvantage more broadly (e.g., income without applying a threshold).

Amongst evidence in the last 10 years, we found one review (Black et al., 2017) showing the quantitative evidence and mechanistic understanding on how child poverty leads to worse early childhood developmental outcomes. There were several primary studies (five cross-sectional, five longitudinal) examining evidence of associations between poverty and a range of developmental outcomes. Only four of these studies examined outcomes in children under 3 years old. Studies in high income countries were predominantly carried out in the US. Two studies used recent national survey data (China 2020, Ecuador 2018/19) while the rest used birth cohort data on children born 1 to 2 decades ago. There was one randomised controlled trial protocol on poverty reduction using unconditional cash gifts on child development in the US. There is no recent evidence in the UK context on how child poverty has impacted early child development.

*Children services in the UK and ECD*

Initial evaluations of Sure Start in 2008 found improved socioemotional development at 3 years, but no effects on educational and socioemotional outcomes at 5 years.^17,18^ However recent evaluations have revealed benefits for school readiness at 5 years in multiple developmental domains for children from low-income families, and improved communication and language regardless of disadvantage. More recent evaluations have focused on longer term benefits for health in adulthood, educational, labour market and welfare outcomes,^19–21^ as well as intergenerational effects.^22^ A recent systematic review of Sure Start showed evidence of benefits on physical health, social functioning and neurodevelopmental difficulties, from Sure Start core services usage, health support worker programmes, health education, parenting interventions. Only one other study assessed the impact of cuts to services on childhood obesity.^23^ In summary, benefits of children services on children’s development has taken years to become evident, and during this time there has been significant cuts to services. There has still not been any evidence quantifying how cuts to services had impacted ECD.

**Added value of this study**

This study provides recent national-level evidence on how geographical and temporal trends in child poverty and under-5 children services spending are associated with ECD in England. We use high quality population level ECD data, providing evidence on the importance of such data in providing rapid intelligence in the UK context to guide the policy decisions targeted at improving ECD.

**Implications of all the available evidence**

Reducing child poverty and increasing spending on under-5 children’s services are key measures to address worsening ECD outcomes since the COVID-19 pandemic and persistent inequalities in ECD.

## Methods

### Data sources and measures

We obtained ECD data between 2017/18 to 2023/24 from the Health Visitor Service Delivery Metrics, published by the Office of Health Improvement and Disparities.^24^ ECD was measured using the Ages and Stages Questionnaire 3 (ASQ-3) at 2 to 2.5 years as part of a universal developmental review.^25^ Our primary outcome was the rate per 100 children *not* developmentally on track in any of the developmental domains assessed on the ASQ-3 (Communication, Gross Motor, Fine Motor, Problem Solving and Personal Social Skills).

Our secondary outcomes were the rate per 100 children not on track in each of the five domains. We excluded LAs with irreconcilable boundary changes (Cumberland, Westmorland and Furness), combined LAs with small population sizes (Isles of Scilly with Cornwall, City of London with Hackney) and focused on data with at least 75% ASQ coverage (See Appendix S2 and Table S1). Since each period corresponded to children born at least 2 years before, the data were for children born between 2016/17 to 2021/22, henceforth 2017 to 2022.

Our exposures were child poverty and under-5 children services spending. Data on child poverty for each local authority was obtained from the Children in Low Income Families dataset, measured as the percentage of children aged 0 to 15 living in households below 60% of OECD median household income, before housing costs.^26^ Data on under-5 children services spending was obtained from Department for Education Local Authority and school expenditure statistics. We used the total gross spending on Sure Start children’s centres and other services for children under 5.^27^ Spending was converted to £ per child aged under five, using midyear population estimates.^28^

For describing variation between local authorities, we also used quintiles of the Income Deprivation Affecting Children Index (IDACI) Average Score from the 2019 Indices of Multiple Deprivation^29^ and region (Yorkshire and The Humber, West Midlands, South West, South East, North West, North East, London, East of England, East Midland).

We controlled for several covariates based on a logic model (Appendix S3), including ASQ coverage, ethnic composition, birth year and excess mortality. ASQ coverage (percentage of eligible 2-to 2.5-year-old children who received an ASQ review) affects the denominator (number of children assessed) in the calculation of rates, so could bias the results if child poverty or free school meal eligibility also influences ASQ coverage. Ethnic composition (Percentage of children aged 0 to 15 of White British/Irish ethnicity based on the 2021 UK Census^30^) was controlled for because some local authorities may be more income deprived due to their ethnic composition and there are known ethnic inequalities in early childhood development. Birth year was intended to capture annual trends and national shocks including due to the COVID-19 pandemic. In addition, we used data on excess mortality during the period of lockdown restrictions (October 2020 and December 2020), from the Office for Health Improvement and Disparities,^31^ to capture residual variation in the COVID-19 shock experienced by different LAs, that was not captured by birth year.

## Statistical analyses

Statistical analyses were carried out in R (V4.3.2). We described variation between local authorities in the exposures and outcomes by birth year, IMD quintile and region, using means and standard deviations; and variation within local authorities (time trends) by IMD quintile and region. We obtained non-parametric Kendall Tau correlations between each exposure.

We used within-between models to estimate associations of child poverty and under-5 spending on ECD. Within-between models enable “within-LA” estimates capturing the extent that annual changes in the exposure is associated with annual changes in the outcome, and “between-LA” estimates capturing the extent that differences between-LA in the exposure is associated with the outcome. Within-LA associations are more robust to confounding by LA characteristics that remain relatively constant over time, but can still be affected by variables that change over time, such as data quality. All models controlled for the covariates described above to address confounding and bias. Outcomes were log-transformed due to positive skew and violation of the assumption of heteroscedasticity in models of the outcome on the original scale. Exposures were scaled to increased interpretability and model convergence.

Exponentiated model coefficients therefore captured the relative change in rates of ECD, for a 10 percentage-point (pp) change in child poverty or free school meal eligibility, or a £100 per child change in annual children spending. Models were run for the rate not on track in any domain (primary outcome), and then by domain (secondary outcomes).

First, we estimated the independent associations between child poverty and under-5 spending on ECD, controlling for covariates. This estimates the detrimental association of child poverty on ECD, after accounting for the co-occurring beneficial association of under-5 spending, and vice-versa. We considered interaction effects between child poverty and under-5 spending.

Second, we re-ran models of child poverty on ECD, before adjusting for under-5 spending and compared this to the estimates adjusted for under-5 spending. This quantifies the extent that under-5 spending attenuated geographical inequalities in ECD or altered the effects of any changes in child poverty on ECD. We used likelihood ratio tests and R^2^ values to compare models of child poverty on ECD with versus without under-5 spending.

## Results

### Variation in child poverty, under-5 spending and ECD

On average across English local authorities, child poverty rose by 3.5 percentage points from 21.5% [95%CI: 20.3; 22.7] in 2017 to 25.0% [23.5; 26.4] in 2022. Under-5 children services spending per child under 5 fell from £242 [216; 219] in 2017 to £187 [157; 216] in 2022 (**Error! Reference source not found**. and Table S2).

**Figure 1.**
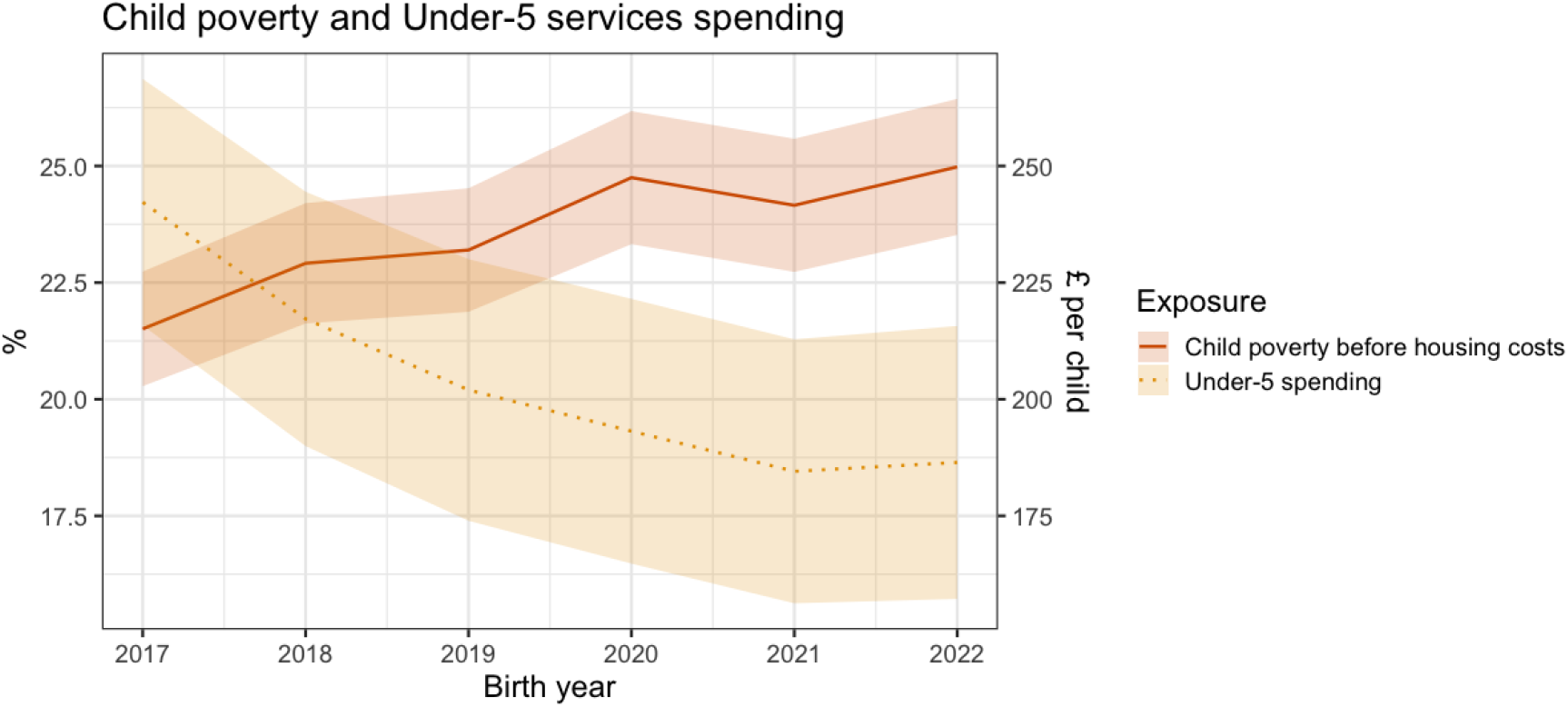
Time trends in child poverty before housing costs and Under-5 services spending

As expected, child poverty was higher in more deprived local authorities (lower IMD quintile) (**Error! Reference source not found**. and Table S3). Under-5 services spending was highest in the most deprived areas (Quintile 1: £290 [254; 327]), and lowest in the least deprived areas (Quintile 5: £151[138; 165]), with a high degree of overlap in the confidence intervals for spending in Quintile 2 and 3. Spending was weakly correlated with child poverty – areas with greater spending had higher percentages of child poverty (Tau, range: 0.15 to 0.17, p<0.001, Table S4).

**Figure 2.**
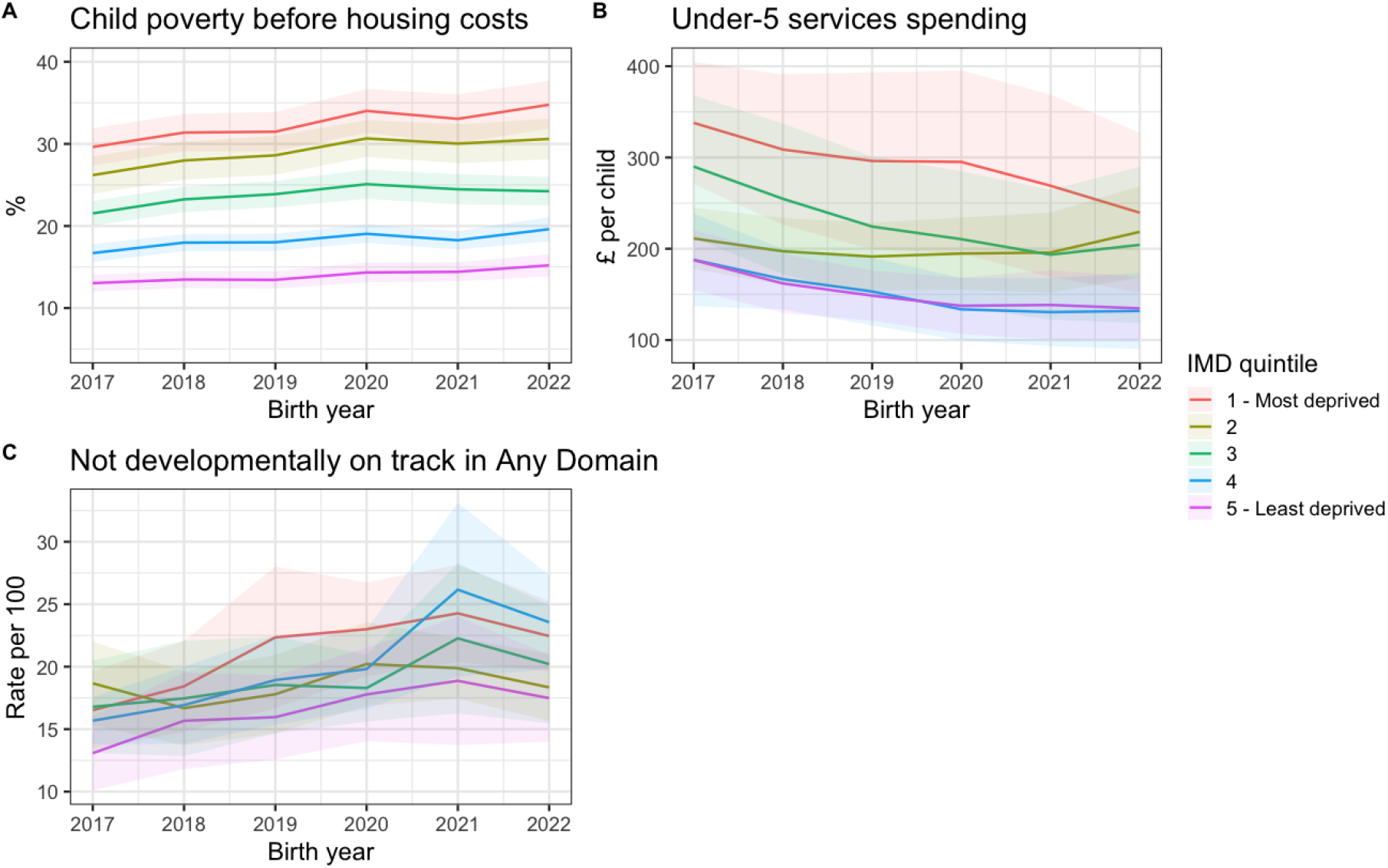
Variation in child poverty and under-5 children’s services spending, and not being on track in Any Domain by IMD 2019 quintile

The rate per 100 children not developmentally on track in Any Domain at 2 to 2.5 years increased for children born in 2017 (15.6 [14.2; 16.9])) to 2020 (19.7 [18.0; 21.5]) and remained stable for children born between 2020 to 2022 (Table S5). Similar time trends were observed for outcomes in each domain. There is a clear inequality gap in not being on track in Any Domain between the most deprived (Quintile 1: 21.5 [19.9; 23.1]) and least deprived quintile (Quintile 5: 16.6 [15.0; 18.1]). However, there was a high degree of overlap in the confidence intervals for local authorities in all quintiles (**Error! Reference source not found**. and Table S6).

Summary trends of the geographic variation in child poverty, under-5 services spending, and ECD comparing local authorities (Figure S1 and Table S7) indicate variation in the outcomes and exposures, both between and within-LA. Within-LA changes in child poverty were relatively small compared to the extent of between-LA variation.

### Association of child poverty and under-5 spending on ECD

Independent of spending and covariates, within-LA annual changes in child poverty were not associated with annual changes in ECD in Any Domain, or in each of the five domains. Between LAs, a 10pp higher child poverty rate between LAs was associated with a 9%(1.09[95%CI: 1.01; 1.18], p=0.031) greater rate of children not on track in Any Domain (Table 1). Child poverty was associated with greater rates of not being on track in all domains except Gross Motor (1.07[1.06; 1.21], p=0.291). The largest association was in Fine Motor (1.25[1.07; 1.45], p=0.004), and similar associations were found in Communication, Problem Solving and Personal Social Skills (Range: 12% to 14%).

**Table 1.** Within-between model of the independent association of child poverty BHC and under-5 children services spending on rates of children not on track in any domain, and in five ECD domains.

|  | Any domain | Communication | Gross Motor | Fine Motor | Problem Solving | Personal Social Skills |
| --- | --- | --- | --- | --- | --- | --- |
| **Within local authority annual change** |  |  |  |  |  |  |
| 10pp annual increase in child poverty | 1 | 0.95 | 0.95 | 0.82 | 0.97 | 1 |
|  | (0.90 – 1.12) | (0.83 – 1.08) | (0.78 – 1.16) | (0.65 – 1.04) | (0.81 – 1.15) | (0.85 – 1.17) |
| £100 per child increase in under-5 spending | **0.96 \*** | 1.01 | 0.99 | **0.94 #** | 0.97 | 0.98 |
|  | **(0.93 – 1.00)** | (0.97 – 1.05) | (0.93 – 1.06) | **(0.87 – 1.01)** | (0.92 – 1.03) | (0.93 – 1.03) |
| **Between local authority difference** |  |  |  |  |  |  |
| 10pp higher rate of child poverty | **1.09 \*** | **1.12 \*\*** | 1.07 | **1.25 \*\*** | **1.14 \*** | **1.14 \*** |
|  | **(1.01 – 1.18)** | **(1.03 – 1.22)** | (0.94 – 1.21) | **(1.07 – 1.45)** | **(1.02 – 1.27)** | **(1.03 – 1.27)** |
| £100 per child higher under-5 spending | **0.97 #** | 0.97 | 0.97 | **0.91 \*\*** | 0.96 | **0.95 \*** |
|  | **(0.93 – 1.00)** | (0.94 – 1.01) | (0.91 – 1.03) | **(0.85 – 0.98)** | (0.91 – 1.01) | **(0.90 – 1.00)** |

Independent of child poverty and covariates, every £100 per child annual increase in spending on under 5 children services was associated with 4% (0.96[0.93; 1.00], p=0.042), lower rate of children not on track in Any Domain. Within-LA associations were stronger for Fine Motor (0.94[0.87; 1.01], p=0.079) compared to other domains. Similar estimates were found for between-LA associations of under-5 spending on Any Domain. Areas with greater under-5 spending had lower rates of children not on track in Fine Motor and Personal Social Skills.

There was no evidence of interaction effects between child poverty and under-5 spending (Table S8).

**Figure 3.**
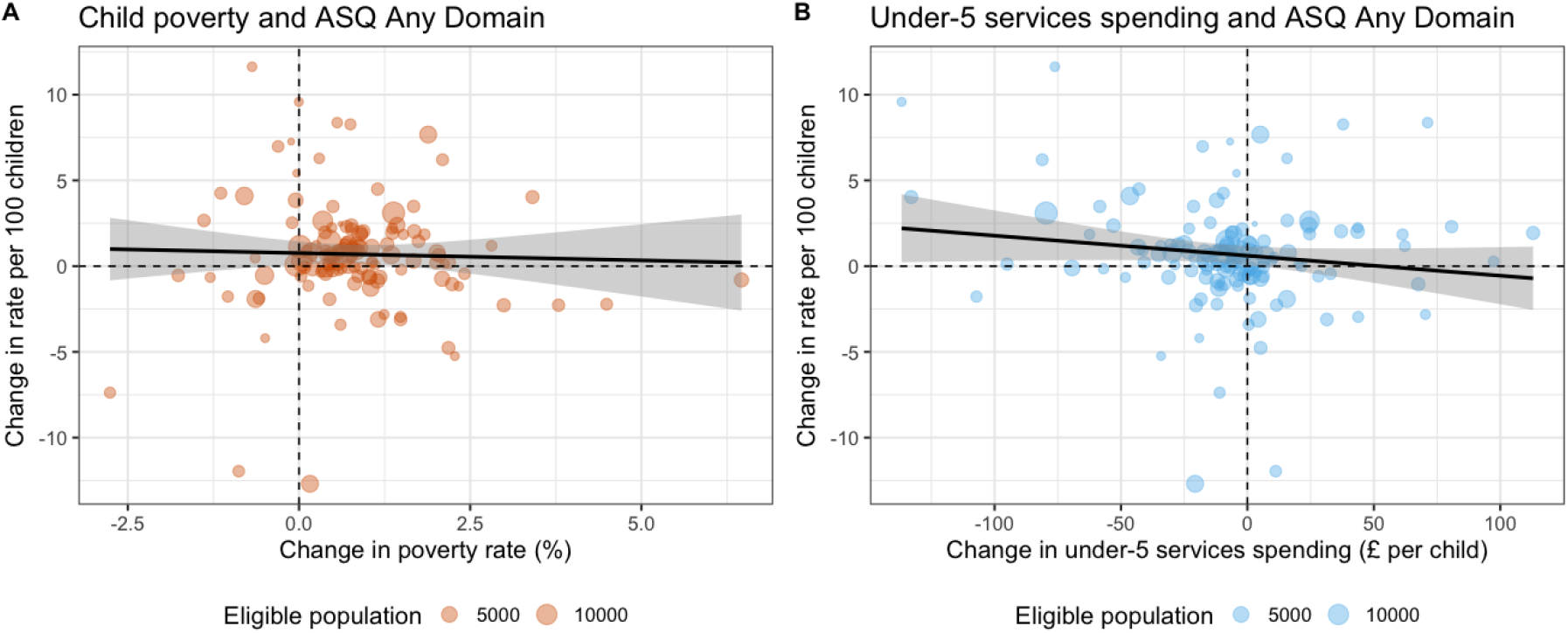
A: Annual changes in child poverty rate and rate of children not on track in development, B: Annual changes in under-5 children services spending and rate of children not on track in development

### Association of child poverty before and after controlling for under-5 spending

Within-LA associations of child poverty on all ECD outcomes remained unchanged whether controlling for children spending or not (Table S9). Between-LA estimates before adjusting for children spending captures the benefits of children spending. For any domain between LA estimates were 2% lower than the model adjusted for under-5 spending (1.07[1.01; 1.16] p=0.076 versus 1.09[1.01; 1.18] p=0.031, Likelihood ratio test: χ^2^ =7.3, p=0.026). This indicates that under-5 spending had attenuated the association of child poverty on ECD by 2 percentage points. When looking at outcomes by domain, the largest attenuation in the association of child poverty on ECD by under-5 spending was observed in Fine Motor (1.19[1.03; 1.38] p=0.09, versus 1.25[1.07;1.45] p=0.004, Likelihood ratio test: χ^2^ =9.9, p=0.007).

### Sensitivity analyses

When restricting the data to those based on 90% ASQ coverage estimates of child poverty BHC and under-5 spending on ECD remained robust (Table S13).We repeated the analysis using child poverty after housing costs which also showed within-LA and between-LA variation (Figure S2, Table S9-12). Using child poverty after housing costs obtained from the Local Child Poverty Statistics by the End Child Poverty Coalition^32^, we found stronger between-LA associations of child poverty on all domains (Table S14). Within-LA associations did not change substantially.

Given limited variation in within-LA child poverty variation, we repeated the analyses using free school meal eligibility instead of child poverty to determine the sensitivity of our analyses to within-LA changes in child poverty. Free school meal eligibility (FSM eligibility) is based on low income and receipt of benefits amongst school-aged children. It has risen consistently since 2017/18,^33^ due to eligibility protections for those at risk of losing out on free school meals during major reforms to the UK benefits system (ie., introduction of Universal Credit) (Figure S3). Therefore, the consistent rise in free school meal eligibility since 2017/18 captures the numbers of children falling into income deprivation, amongst those who were not previously income deprived. We used Department for Education schools and pupil characteristics data, on percentage of pupils in state-funded primary schools (5 to 11 years old) meeting eligibility for Free School Meals in each local authority.^33^

We focus on within-LA associations here, but briefly, between-LA associations of FSM eligibility on ECD showed similar but less precise associations compared to associations of child poverty on ECD (Table S15). Within-LA associations indicated that for a 10pp increase in FSM eligibility, the rate of children not on track in Any Domain increased by 16% (1.16[1.00; 1.36]). Positive but imprecise associations were observed in all individual domains (e.g., Fine Motor: 1.19[0.87; 1.65]; Problem Solving: 1.35[1.06; 1.71]) (Table S15). Within-LA associations of under-5 spending attenuated the association of FSM eligibility on rate of children not on track in Any Domain (1.14[0.98; 1.35], χ^2^ =8.0, p=0.018), indicating that increases within-LA in under-5 spending were positively correlated with FSM eligibility and buffered the association of FSM eligibility on ECD.

**Figure 4.**
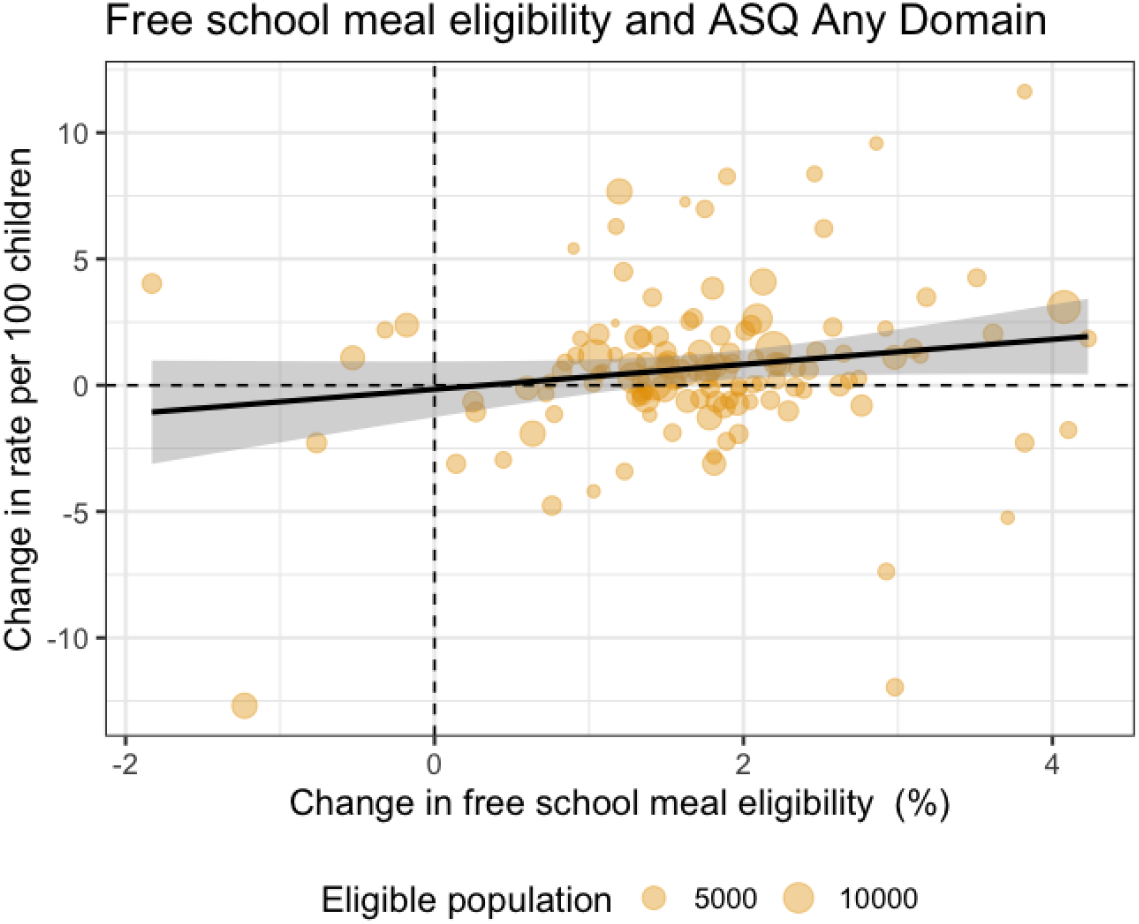
Annual changes in free school meal eligibility in state-funded primary schools and rate of children not on track in development

## Discussion

This was an area-level study of ECD at 2 to 2.5 years in children born between 2017 and 2022. Areas with higher child poverty had higher rates of children not developmentally on track at 2 to 2.5 years, but annual changes in child poverty were not associated with ECD. Independent of the rate of child poverty, greater spending on under-5 services was associated with better ECD outcomes, with consistent within-LA and between-LA estimates. Higher spending in areas with higher poverty attenuated the association of child poverty on ECD, indicating that an area spending more on early years had better ECD outcomes compared to other areas with similar levels of child poverty.

When examining ECD domains, areas with higher child poverty had worse ECD in all domains except Gross Motor. We found stronger evidence for the association between under-5 spending on Fine Motor, over other domains, as well as stronger evidence that spending attenuated the detrimental association of child poverty on ECD for the Fine Motor domain, over other domains.

Our findings capture two key drivers of ECD outcomes: child poverty and spending. This is in line with well-established evidence that poverty is damaging for children’s development.^2^ Our findings also extend the limited evidence base on the early years benefit of spending on Sure Start children centres on socioemotional development at 3 years old,^17^ and obesity and other educational outcomes from 5 years old.^20,23^

Family socioeconomic circumstances in the UK worsened during the COVID-19 pandemic, including financial circumstances, employment uncertainty, isolation, wellbeing, all of which are associated with poverty status and could contribute to worsening ECD outcomes.^34^ As the increase in child poverty during the pandemic was buffered by uplifts to Universal Credit, changes in financial poverty may not fully capture changes in familial socioeconomic context. Reduced social opportunities during the pandemic also contributed to poorer ECD outcomes, making it difficult to separate the associations of child poverty, over and above pandemic-related effects which we controlled for.

Changes in spending likely drive ECD outcomes by affecting the value delivered and affecting the number of children falling through the net. Sure Start was originally developed to have a universal reach, but funding cuts have led to fewer centres, reduced hours, reduced programmes and services on offer, and referral-based access.^14^ This makes it difficult to deliver an accessible service meeting the needs of disadvantaged families.

We show the pervasive impact of child poverty across various developmental domains. Our non-statistically significant finding of child poverty on gross motor development contrasts with an individual-level study.^35^ This is likely because area-level aggregates are diluted.^36^ For under-5 spending, the strongest evidence was its benefits on fine motor development. However, the pandemic had a larger impact on communication, problem solving and personal social skills than motor skills.^5^ Benefits of under-5 spending on these domains may have been obscured by the pandemic, because children simply did not have the opportunities to develop those skills.

### Implications on research, policy and practice

Future research on the long-term impacts of early years services can address criticism that children who did not benefit from early years programmes still catch up in development subsequently.^37^ There is also accumulating evidence of short and long term benefits of Sure Start.^20,21,38^, but a key question is how early developmental benefits attributable to early childhood programs may cascade onto later benefits.

The projected rise in child poverty is expected to implicate several areas of child health,^39^ and early developmental delays are projected to implicate economic productivity.^5^ Alongside poverty-reduction measures, spending more on early childhood services and programs are one way to mitigate the impact of poverty on ECD. Although the cost effectiveness of preschool programs has been questioned, the long term benefits on labour market productivity, crime and costly late interventions, likely far exceeds the costs of preschool programs.^40^

On average, spending was falling and ECD was worsening during this period. Our findings indicate a £100 per child fall in spending (selected to reflect the average range of spending changes during this period) was contributing to a factor of 4% worse ECD outcomes, even after controlling for COVID-19 shocks. Compared to 2010/11, spending per child on Sure Start children services fell from £600 per child to £200 in 2023/24.^41^ This suggests that the latest rate of developmental delays could be 16% lower, if spending was at 2010 levels.

Spending on early years services and reducing child poverty are important policy routes towards achieving school readiness targets. As ECD and school readiness are key to later economic prosperity, investments in improving ECD are likely to be cost effective in the long run. The UK government’s renewed commitment to prevention and to shifting care from hospitals to community settings is a positive step, as is the emphasis on early years provision through Better Start centres, which succeed Sure Start. However, these initiatives must be properly funded, expanded in line with local need, and accompanied by overdue measures to tackle rising child poverty in the UK. Questions remain about whether the budget for Better Start (c. £500million) is sufficient, given it is approximately one fifth of the Sure Start investment.

### Strengths and limitations

The use of within-between models has strengths, as well as limitations. Within-LA estimates are more robust to confounding by variables that differ between LAs, but do not vary significantly over time, unlike for example the number of Sure Start centres. Although there are also within-LA changes over time in the number of Sure Start children centres, closures in Sure Start children centres were relatively stable by the start of the study period.^42^ Varying ASQ coverage could also influence within-LA estimates, but we controlled for this. The consistent within-LA and between-LA estimates demonstrates strong evidence that greater under-5 services spending improves ECD.

Within-LA variation in child poverty during this period was noisy due to the introduction of universal credit, and temporary Universal Credit uplifts during the pandemic. With relatively few timepoints, having sufficient within-LA variation is also important for a well-powered analysis. Previous work over a longer period, which saw larger changes in child poverty, have found robust associations of child poverty on child health outcomes^43,44^. To address this limitation, we considered a related measure, FSM eligibility. However, estimates were less precise because FSM eligibility misses as many as a third of children in poverty.

## Conclusions

Area-level differences in child poverty across the UK drives inequalities in ECD, as early as 2 to 2.5 years, but spending on children services reduced these inequalities. Alongside the effects of COVID-19, the overall fall in spending during this period contributed to worsening ECD outcomes. The alarming projections of what child poverty would look like in the UK underscores the importance of investment in the early years to alleviate the detrimental effects of poverty on ECD.

## Data Availability

All data produced in the present work are contained in the manuscript.

## Acknowledgements

YWC is part-funded by an NIHR Research Professorship (NIHR302438) awarded to DTR and the NIHR Oxford Health Biomedical Research Consortium. CM was funded by an NIHR Undergraduate Internship Programme (NIHR304390), awarded to YWC. DTR, LA and DB are funded by the NIHR School for Public Health Research (grant reference number NIHR204000). DTR is also funded on an NIHR Research Professorship (NIHR302438). LM is funded by NIHR Applied Research Collaboration Greater Manchester (ARC-GM; NIHR200174). The views expressed are those of the authors and not necessarily those of the NIHR or the Department of Health and Social Care. Hannah Mohammad contributed to screening hits from the literature search and extracting data presented in the Research in Context box. For the purpose of open access, the author has applied a Creative Commons Attribution (CC BY) licence to any Author Accepted Manuscript version arising from this submission.

## Notes

### Competing Interest Statement

The authors have declared no competing interest.

### Author Declarations

The study used only openly available human data that were originally located at: https://www.gov.uk/government/statistics/child-development-outcomes-at-2-to-2-and-a-half-years-quarterly-data-for-2021-to-2022; https://www.gov.uk/government/collections/children-in-low-income-families-local-area-statistics https://explore-education-statistics.service.gov.uk/find-statistics/la-and-school-expenditure https://www.nomisweb.co.uk/datasets/pestsyoala https://opendatacommunities.org/resource?uri=http%3A%2F%2Fopendatacommunities.org%2Fdata%2Fsocietal-wellbeing%2Fimd2019%2Findicesbyla https://www.ons.gov.uk/peoplepopulationandcommunity/culturalidentity/ethnicity/bulletins/ethnicgroupenglandandwales/census2021 https://www.lboro.ac.uk/research/crsp/our-research/local-child-poverty-indicators/

