## Supplemental Figures, Tables and Appendices for "Child poverty and under-5 children services spending on early childhood development in England – a longitudinal local area ecological study of children born between 2017 and 2022"

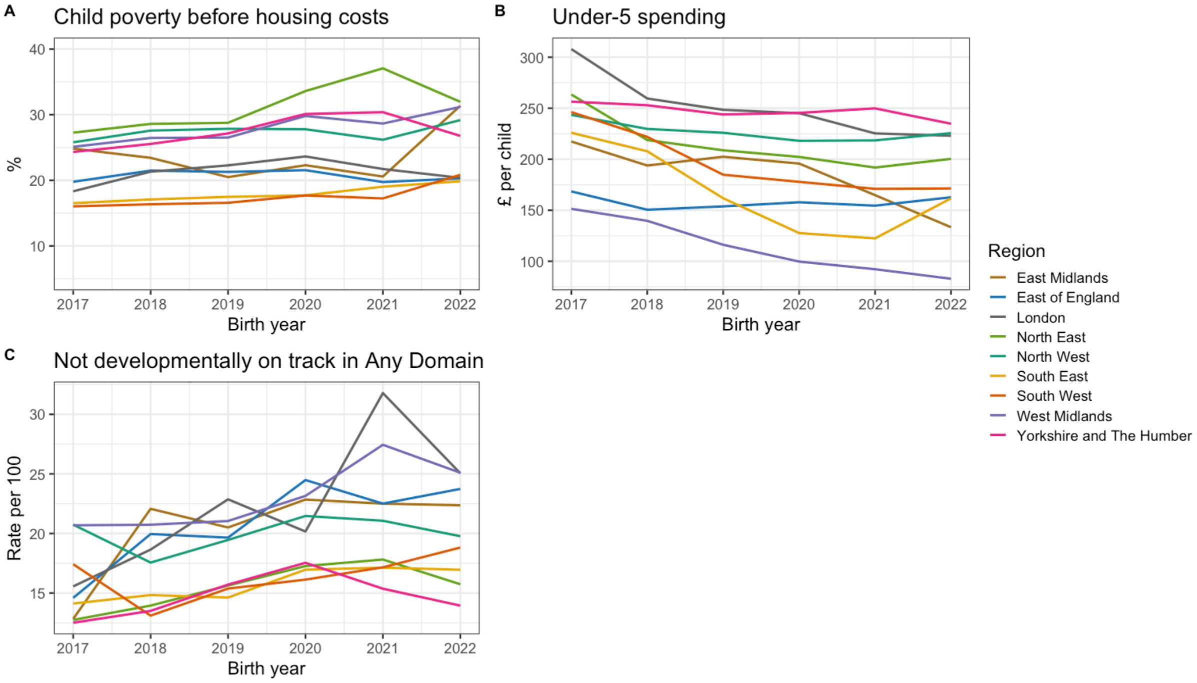


**Figure S1. Percentage child poverty before housing costs (BHC), £ per child spending on under-5 children services, and rate per 100 children not on track in any domain by region of England, capturing within-LA and between-LA variation in the exposures and outcome.**

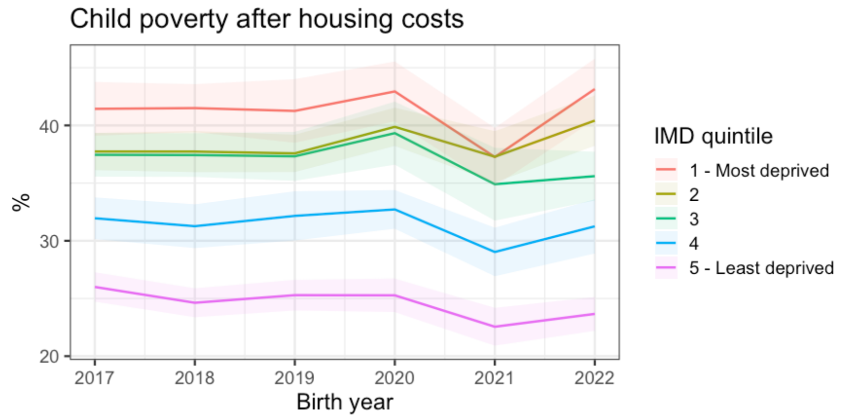


**Figure S2. Time trends by IMD2019 in Child Poverty AHC. Within-LA changes showed sharp changes during and after the COVID-19 pandemic due to temporary universal credit uplifts that bolstered family incomes.**

A

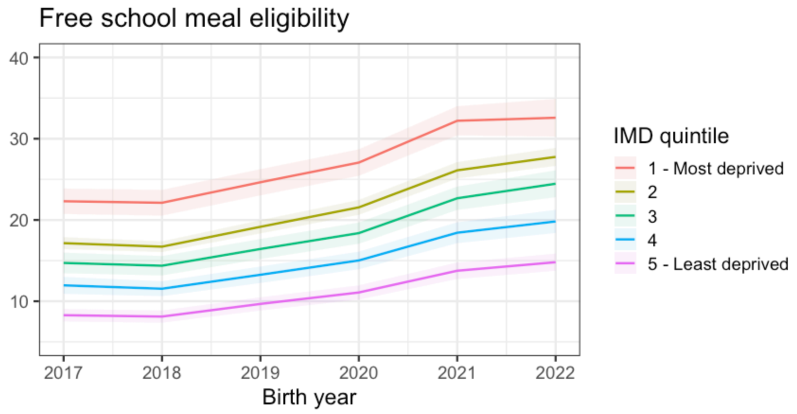


B

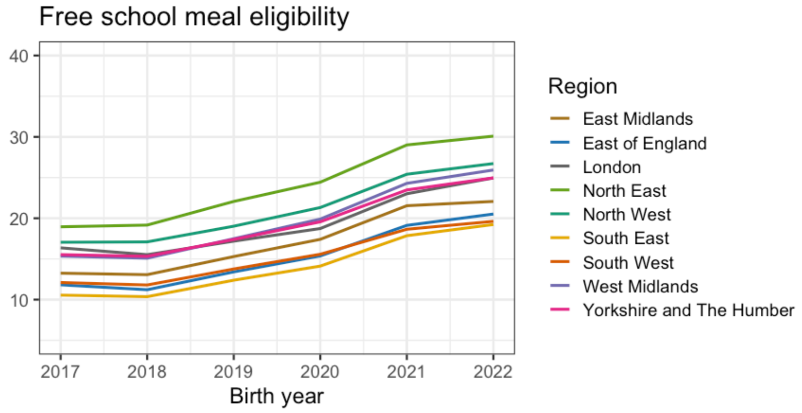


**Figure S3. Percentage Free school meal eligibility by IMD 2019 quintile and region of England, from 2017 to 2022**

### Supplemental Tables

**Table S1. Analysis sample – number (%) of LAs with above 75% coverage and publishable data on ASQ domains.** Only 3 datapoints from 2017 were excluded due to incompleteness of data on children services spending and free school meal eligibility.

| Birth year | Any domain | Communication | Gross Motor | Fine Motor | Problem Solving | Personal Social Skills |
|---|---|---|---|---|---|---|
| 2017 | 103 (69.1%) | 106 (71.1%) | 106 (71.1%) | 106 (71.1%) | 106 (71.1%) | 106 (71.1%) |
| 2018 | 119 (79.9%) | 119 (79.9%) | 119 (79.9%) | 119 (79.9%) | 119 (79.9%) | 119 (79.9%) |
| 2019 | 101 (67.8%) | 101 (67.8%) | 101 (67.8%) | 101 (67.8%) | 101 (67.8%) | 101 (67.8%) |
| 2020 | 112 (75.2%) | 112 (75.2%) | 112 (75.2%) | 112 (75.2%) | 111 (74.5%) | 112 (75.2%) |
| 2021 | 106 (71.1%) | 106 (71.1%) | 106 (71.1%) | 106 (71.1%) | 106 (71.1%) | 106 (71.1%) |
| 2022 | 118 (79.2%) | 118 (79.2%) | 118 (79.2%) | 118 (79.2%) | 118 (79.2%) | 118 (79.2%) |
| Total | 659 | 662 | 662 | 662 | 661 | 662 |

**Table S2. Mean (95%CI) annual trends in child poverty before housing costs (BHC) and under-5 children services spending in English LAs.**

| Birth year | Child poverty BHC | Under-5 spending |
|---|---|---|
| 2017 | 21.5% (20.3 to 22.7) | £242 (216 to 269) |
| 2018 | 22.9% (21.6 to 24.2) | £217 (190 to 244) |
| 2019 | 23.2% (21.9 to 24.5) | £202 (174 to 230) |
| 2020 | 24.8% (23.3 to 26.2) | £193 (165 to 222) |
| 2021 | 24.2% (22.7 to 25.6) | £185 (156 to 213) |
| 2022 | 25.0% (23.5 to 26.4) | £186 (157 to 216) |

**Table S3. Mean (95%CI) percentage child poverty BHC and £ per child under-5 children services spending in English LAs by IMD 2019 quintile**

| IMD 2019 | Child poverty BHC | Under-5 spending |
|---|---|---|
| 1 - Most deprived | 32.4% (31.3 to 33.5) | £290 (254 to 327) |
| 2 | 29.0% (28.0 to 30.0) | £202 (185 to 218) |
| 3 | 23.7% (23.0 to 24.4) | £230 (198 to 261) |
| 4 | 18.3% (17.8 to 18.8) | £150 (134 to 167) |
| 5 - Least deprived | 14.0% (13.5 to 14.5) | £151 (138 to 165) |

**Table S4. Kendall Correlation between percentage child poverty BHC, £ per child spending on under-5 children services by birth year**

| Birth year | Kendall Tau | p |
|---|---|---|
| 2017 | 0.15 | 0.006 |
| 2018 | 0.16 | 0.003 |
| 2019 | 0.20 | 0.000 |
| 2020 | 0.21 | 0.000 |
| 2021 | 0.17 | 0.002 |
| 2022 | 0.17 | 0.002 |

**Table S5. Mean (95%CI) annual trends in rate per 100 children not on track in Any Domain and in each of the five ASQ domains**

| Birth year | Any domain | Communication | Gross Motor | Fine Motor | Problem Solving | Personal Social Skills |
|---|---|---|---|---|---|---|
| 2017 | 15.6 (14.2 to 16.9) | 10 (9.1 to 10.8) | 6.1 (5.2 to 6.9) | 5.9 (4.9 to 6.9) | 5.5 (4.7 to 6.3) | 6.6 (5.7 to 7.5) |
| 2018 | 16.4 (14.9 to 17.9) | 11 (9.9 to 12.1) | 5.8 (4.9 to 6.6) | 5.6 (4.6 to 6.6) | 5.9 (5 to 6.8) | 6.6 (5.8 to 7.5) |
| 2019 | 17.6 (15.6 to 19.6) | 12.7 (10.9 to 14.4) | 7 (5.5 to 8.6) | 6.8 (5.1 to 8.4) | 7 (5.4 to 8.5) | 8.2 (6.6 to 9.7) |
| 2020 | 19.7 (18 to 21.5) | 14.5 (13.1 to 15.9) | 7 (5.9 to 8.1) | 7.3 (5.9 to 8.7) | 8 (6.8 to 9.2) | 9.5 (8.3 to 10.7) |
| 2021 | 20.6 (18.9 to 22.4) | 14.7 (13.1 to 16.2) | 7.8 (6.5 to 9.1) | 7.8 (6.4 to 9.2) | 8.8 (7.5 to 10) | 10.1 (8.8 to 11.5 ) |
| 2022 | 19.8 (18.2 to 21.4) | 13.7 (12.5 to 14.9) | 6.6 (5.9 to 7.4) | 6.8 (5.8 to 7.8) | 7.9 (7 to 8.7) | 9.0 (8.1 to 9.9) |

#### Table S6. Mean (95%CI) rate per 100 children not on track in Any Domain and in each of the five ASQ domains, by IMD 2019 quintile (1-Most deprived, 5-Least deprived)

| IMD 2019 | Any domain | Communication | Gross Motor | Fine Motor | Problem Solving | Personal Social Skills |
|---|---|---|---|---|---|---|
| 1 | 21.5 (19.9 to 23.1) | 15.5 (14.2 to 16.9) | 8.4 (7 to 9.7) | 9.7 (8.1 to 11.3) | 9.2 (7.9 to 10.6) | 10.8 (9.2 to 12.4) |
| 2 | 18.6 (17.4 to 19.8) | 13.2 (12.3 to 14.1) | 6.2 (5.5 to 6.9) | 6.5 (5.6 to 7.3) | 7.4 (6.6 to 8.1) | 8.1 (7.4 to 8.9) |
| 3 | 19.1 (17.2 to 20.9) | 13.8 (12 to 15.5) | 5.9 (5 to 6.7) | 6.2 (5.2 to 7.3) | 7 (6.2 to 7.9) | 7.9 (7 to 8.8) |
| 4 | 20.1 (18.4 to 21.8) | 14.1 (12.5 to 15.8) | 8 (6.6 to 9.3) | 7.2 (5.8 to 8.6) | 7.9 (6.4 to 9.3) | 9.4 (7.9 to 10.8) |
| 5 | 16.6 (15 to 18.1) | 11.3 (10 to 12.7) | 7 (6.1 to 7.9) | 6.2 (5.2 to 7.1) | 7.1 (6.1 to 8) | 8.1 (7.1 to 9.1) |

#### Table S7. Mean (95%CI) percentage child poverty BHC, £ per child spending on under-5 children services, and rate per 100 children not on track in any domain in English LAs by region of England.

| Region | Child poverty BHC | Under-5 spending | ASQ Any domain |
|---|---|---|---|
| East Midlands | 23.8 (21.5 to 26.1) | 154 (130 to 178) | 21.9 (20.3 to 23.5) |
| East of England | 20.7 (19.1 to 22.2) | 158 (145 to 171) | 20.8 (18.3 to 23.3) |
| London | 21.3 (20.4 to 22.2) | 252 (215 to 288) | 22.7 (21.2 to 24.2) |
| North East | 31.2 (30.1 to 32.3) | 214 (178 to 250) | 15.6 (14 to 17.2) |
| North West | 27.4 (25.9 to 28.9) | 227 (211 to 243) | 20.2 (19 to 21.3) |
| South East | 17.9 (16.8 to 19.1) | 168 (149 to 186) | 15.9 (14.9 to 17) |
| South West | 17.5 (16.7 to 18.3) | 191 (172 to 210) | 16.3 (15.5 to 17.1) |
| West Midlands | 28 (26 to 29.9) | 114 (99 to 129) | 22.9 (21 to 24.8) |
| Yorkshire and The Humber | 27.4 (25.9 to 28.9) | 247 (213 to 282) | 14.7 (14 to 15.5) |

#### Table S8. Within-between model (with interaction effects) of the independent association of child poverty BHC and under-5 children services spending on rates of children not on track in any domain, and in five ECD domains.

| | Any domain | Communication | Gross Motor | Fine Motor | Problem Solving | Personal Social Skills |
|---|---|---|---|---|---|---|
| **Within local authority annual change** | | | | | | |
| 10pp annual increase in child poverty | 1.02 | 0.99 | 0.9 | 0.88 | 0.98 | 1 |
| | (0.90 – 1.16) | (0.85 – 1.16) | (0.71 – 1.13) | (0.68 – 1.15) | (0.80 – 1.20) | (0.83 – 1.20) |
| £100 per child increase in under-5 spending | 0.96 | 0.99 | 0.93 | 1.02 | 0.94 | 0.94 |
| | (0.87 – 1.06) | (0.88 – 1.12) | (0.77 – 1.11) | (0.83 – 1.26) | (0.81 – 1.10) | (0.81 – 1.08) |
| Poverty x Spending | 1 | 1 | 1.02 | 0.97 | 1.01 | 1.02 |
| | (0.97 – 1.03) | (0.97 – 1.05) | (0.96 – 1.09) | (0.91 – 1.04) | (0.96 – 1.06) | (0.97 – 1.07) |
| **Between local authority difference** | | | | | | |
| 10pp higher rate of child poverty | 1.13 | 1.16 | 1.09 | 1.27 | 1.13 | 1.13 |
| | (0.98 – 1.31) | (0.99 – 1.35) | (0.86 – 1.39) | (0.96 – 1.68) | (0.92 – 1.39) | (0.93 – 1.37) |
| £100 per child higher under-5 spending | 1.02 | 1.01 | 1 | 0.93 | 0.95 | 0.93 |

| | | | | | | |
|---|---|---|---|---|---|---|
| | (0.85 – 1.23) | (0.84 – 1.23) | (0.75 – 1.34) | (0.66 – 1.31) | (0.74 – 1.22) | (0.73 – 1.18) |
| Poverty x Spending | 0.98 | 0.98 | 0.99 | 0.99 | 1 | 1.01 |
| | (0.91 – 1.05) | (0.92 – 1.06) | (0.88 – 1.10) | (0.87 – 1.13) | (0.91 – 1.11) | (0.92 – 1.10) |

*Note: All models controlled for Year, Ethnicity, ASQ coverage, and COVID-19 mortality*

#### Table S9. Between and within-LA associations of child poverty, not adjusted for under-5 children services spending and likelihood ratio tests for adjusted versus unadjusted models.

| | Any domain | Communication | Gross Motor | Fine Motor | Problem Solving | Personal Social Skills |
|---|---|---|---|---|---|---|
| **Between local authority difference, not adjusted for under-5 spending** | | | | | | |
| 10pp higher rate of child poverty | 1.07 | **1.11** * | 1.06 | **1.19** * | **1.12** * | **1.12** * |
| | (0.99 – 1.16) | **(1.02 – 1.20)** | (0.93 – 1.19) | **(1.03 – 1.38)** | **(1.00 – 1.24)** | **(1.01 – 1.24)** |
| **Within local authority annual change, not adjusted for under-5 spending** | | | | | | |
| 10pp higher rate of child poverty | 1 | 0.95 | 0.95 | 0.82 | 0.96 | 0.99 |
| | (0.90 – 1.12) | (0.83 – 1.08) | (0.78 – 1.16) | (0.65 – 1.03) | (0.81 – 1.15) | (0.85 – 1.17) |
| **Likelihood ratio test (adjusted vs unadjusted for under-5 spending** | | | | | | |
| Chi(df=2) | 7.3 | 2.2 | 1.0 | 9.9 | 3.6 | 5.0 |
| p | 0.026 | 0.340 | 0.619 | 0.007 | 0.165 | 0.082 |

#### Table S10. Mean (95%CI) annual trends in exposures used in sensitivity analyses – percentage Child poverty after housing costs (AHC) and Free school meal eligibility

| Year | Child poverty AHC | Free school meal eligibility |
|---|---|---|
| 2017 | 35.0 (33.9 to 36.2) | 14.9 (14.0 to 15.7) |
| 2018 | 34.6 (33.4 to 35.9) | 14.5 (13.7 to 15.4) |
| 2019 | 34.8 (33.6 to 36.1) | 16.6 (15.7 to 17.6) |
| 2020 | 36.2 (34.8 to 37.5) | 18.6 (17.6 to 19.6) |
| 2021 | 32.4 (31.0 to 33.7) | 22.6 (21.4 to 23.7) |
| 2022 | 35 (33.5 to 36.4) | 24 (22.8 to 25.2) |

#### Table S11. Mean (95%CI) in exposures used in sensitivity analyses by IMD 2019 quintile - percentage Child poverty AHC and Free school meal eligibility

| IMD 2019 | Child poverty AHC | Free school meal eligibility |
|---|---|---|
| 1 - Most deprived | 41.3 (40.2 to 42.3) | 26.9 (25.9 to 27.9) |
| 2 | 38.4 (37.5 to 39.4) | 21.4 (20.7 to 22.1) |
| 3 | 37 (36.3 to 37.7) | 18.5 (17.7 to 19.3) |
| 4 | 31.4 (30.9 to 31.9) | 15.1 (14.4 to 15.7) |
| 5 - Least deprived | 24.6 (24.1 to 25) | 11 (10.4 to 11.5) |

#### Table S12. Mean (95%CI) in exposures used in sensitivity analyses by region of England – percentage Child poverty AHC and Free school meal eligibility

| Region | Child poverty AHC | Free school meal eligibility |
|---|---|---|
| East Midlands | 30.3 (29 to 31.7) | 17.2 (15.8 to 18.6) |
| East of England | 31.1 (29.6 to 32.7) | 15.2 (14.5 to 16) |
| London | 39.4 (38.2 to 40.6) | 19.3 (18.3 to 20.2) |
| North East | 39.8 (39.1 to 40.5) | 24 (22.9 to 25) |
| North West | 36 (35 to 37) | 21.1 (20.1 to 22.1) |
| South East | 28.3 (27.1 to 29.4) | 14.1 (13.1 to 15.1) |
| South West | 27.9 (27.1 to 28.8) | 15.4 (14.6 to 16.3) |

| | | |
|---|---|---|
| West Midlands | 38.9 (37.7 to 40.1) | 19.7 (18.3 to 21) |
| Yorkshire and The Humber | 34.4 (33.4 to 35.3) | 19.4 (18.5 to 20.2) |

**Table S13.** Within-between model of the independent association of child poverty BHC and under-5 children services spending on rates of children not on track in any domain, and in five ECD domains.

| | **Any domain** | **Communication** | **Gross Motor** | **Fine Motor** | **Problem Solving** | **Personal Social Skills** |
|---|---|---|---|---|---|---|
| **Within local authority annual change** | | | | | | |
| 10pp annual increase in child poverty | 1.03 | 0.98 | 0.92 | 0.93 | 0.98 | 1.01 |
| | (0.91 – 1.16) | (0.85 – 1.12) | (0.74 – 1.13) | (0.73 – 1.18) | (0.81 – 1.18) | (0.85 – 1.21) |
| £100 per child increase in under-5 spending | *0.97 #* | 1.01 | 1 | 0.96 | 0.99 | 0.99 |
| | *(0.93 – 1.00)* | (0.96 – 1.05) | (0.94 – 1.07) | (0.89 – 1.03) | (0.94 – 1.05) | (0.94 – 1.04) |
| **Between local authority difference** | | | | | | |
| 10pp higher rate of child poverty | **1.11** * | **1.15** ** | 1.02 | **1.18** * | **1.13** * | **1.14** * |
| | **(1.02 – 1.20)** | **(1.05 – 1.26)** | (0.89 – 1.17) | **(1.01 – 1.39)** | **(1.00 – 1.27)** | **(1.02 – 1.28)** |
| £100 per child higher under-5 spending | 0.97 | 0.97 | 0.98 | *0.93 #* | 0.96 | *0.95 #* |
| | (0.93 – 1.01) | (0.93 – 1.01) | (0.92 – 1.04) | *(0.87 – 1.00)* | (0.91 – 1.01) | *(0.90 – 1.00)* |

**Table S14.** Within-between model of the independent association of child poverty AHC and under-5 children services spending on rates of children not on track in any domain, and in five ECD domains.

| | **Any domain** | **Communication** | **Gross Motor** | **Fine Motor** | **Problem Solving** | **Personal Social Skills** |
|---|---|---|---|---|---|---|
| **Within local authority annual change** | | | | | | |
| 10pp annual increase in child poverty | 1.01 | 1.03 | 0.98 | **0.85** * | 0.99 | 0.98 |
| | (0.94 – 1.07) | (0.96 – 1.12) | (0.87 – 1.10) | **(0.75 – 0.98)** | (0.89 – 1.09) | (0.89 – 1.07) |
| £100 per child increase in under-5 spending | **0.96** * | 1.01 | 0.99 | 0.93 | 0.97 | 0.98 |
| | **(0.93 – 1.00)** | (0.97 – 1.05) | (0.93 – 1.06) | (0.87 – 1.00) | (0.92 – 1.03) | (0.93 – 1.03) |
| **Between local authority difference** | | | | | | |
| 10pp higher rate of child poverty | **1.11** * | **1.16** ** | 1.13 | **1.35** *** | **1.19** ** | **1.17** * |
| | **(1.01 – 1.22)** | **(1.06 – 1.28)** | (0.98 – 1.31) | **(1.13 – 1.60)** | **(1.05 – 1.35)** | **(1.03 – 1.31)** |
| £100 per child higher under-5 spending | 0.97 | 0.97 | 0.97 | **0.91** ** | 0.96 | **0.95** * |
| | (0.93 – 1.00) | (0.93 – 1.01) | (0.91 – 1.03) | **(0.84 – 0.97)** | (0.91 – 1.01) | **(0.90 – 1.00)** |

Table S15. Within-between model of the independent association of Free School Meal Eligibility and under-5 children services spending on rates of children not on track in any domain, and in five ECD domains.

| FSM eligibility | **Any domain** | **Communication** | **Gross Motor** | **Fine Motor** | **Problem Solving** | **Personal Social Skills** |
|---|---|---|---|---|---|---|
| **Between local authority difference (10pp higher rate)** | | | | | | |
| Adjusted for under-5 spending | 1.08 | 1.13 * | 0.98 | 1.2 | 1.09 | 1.09 |

| | | | | | | |
|---|---|---|---|---|---|---|
| | (0.97 – 1.21) | (1.01 – 1.26) | (0.82 – 1.16) | (0.97 – 1.47) | (0.94 – 1.27) | (0.95 – 1.26) |
| Not adjusted for under-5 services spending | 1.03 | 1.08 | 0.96 | 1.06 | 1.04 | 1.02 |
| | (0.94 – 1.14) | (0.97 – 1.19) | (0.82 – 1.11) | (0.88 – 1.27) | (0.90 – 1.18) | (0.90 – 1.16) |
| **Within local authority difference (10pp annual increase)** | | | | | | |
| Adjusted for under-5 spending | 1.16 # | 1.11 | 1.11 | 1.19 | 1.35 * | 1.21 |
| | (1.00 – 1.36) | (0.93 – 1.33) | (0.84 – 1.46) | (0.87 – 1.64) | (1.06 – 1.71) | (0.97 – 1.51) |
| Not adjusted for under-5 services spnding | 1.14 | 1.11 | 1.1 | 1.16 | **1.33** * | 1.2 |
| | (0.98 – 1.34) | (0.92 – 1.33) | (0.84 – 1.45) | (0.84 – 1.59) | **(1.05 – 1.68)** | (0.96 – 1.50) |
| **Likelihood ratio test (adjusted vs not adjusted for under-5 spending)** | | | | | | |
| Chi(df=2) | 8.0 | 2.8 | 0.3 | 9.7 | 3.5 | 4.3 |
| p | 0.018 | 0.248 | 0.846 | 0.008 | 0.171 | 0.118 |

**Appendix S1. Background of Sure Start**

Sure Start began in 1999 as a multiagency programme to targeted at the most disadvantaged areas based on income deprivation affecting children. It was later extended to all areas from 2003 ("a Children's Centre in every community"), but with lower levels of provision for less deprived areas.[1,2] Between 2003 – 2010, it developed into a one-stop centre (Sure Start Children's Centre) for early support and access to specialised help, linking early education, health and parenting, provisioned by local authorities since 2005.[2] Sure Start takes a two-generation approach focusing on both parents and children.[2] For example, it provides outreach services, home visiting, support for families and parents (including employment support and poverty prevention), preventative services, resources and evidence-based parenting programmes to support child health and development, support for special needs.[2]

Since 2010, there has been no ring-fenced budget for Sure Start as funding was merged with funding for other programmes.[1] From 2010, to manage tightened financial budgets, there was a significant decline in the number of Sure Start centres as well as reorganisation of the services offered.[1,2] From around 3600 centres in 2010, the number of Sure Start centres declined to 3050 by 2019.[3] Although closure of Sure Start centres made news headlines, there was in fact considerable variation in the approach taken by local authorities. An evaluation shows that a minority of local authorities made major reductions to early intervention funding through closures of Sure Start centres. The majority of local authorities maintained the same number of Sure Start centres as 2009, but reorganised service delivery such as cutting back services (full-time to part-time, reduced range of services on offer), and reorganising (e.g. serving a wider age group, linking to another programme). While still available across deprived and non-deprived areas, from 2013 access to Sure Start children centres shifted from universal open access to targeted access based on identified need in high risk families.[1]

Reorganisations and closures of Sure Start children's centres were a reaction to tightened financial budgets. Spending on children's services can be divided into three phases: a fall in spending from 2010 to 2015/16 in response to funding cuts by central government, rise in spending from 2016/17 to 2020/21 due to increased local authority revenues,[4] and a marked increase in spending in 2021/22 compared to the previous year continuing into 2022/23.[5] However, only £1 out of every £5 of increased spending was on early intervention services across children centres, family support and young people services. Despite increases in spending, there was a 71% reduction in real terms of early intervention funding between 2010/11 and 2019/20.[1] Despite marked increases in children spending post-pandemic, in 2021/22, spending on early interventions was just over £2 billion, a 46% reduction of the 2010/11 figure of 3.7 billion.[4] In contrast, spending on late interventions grew from around £6 billion to £8.8 billion between 2010/11 and 2021/22.[4] Regional variation in spending, to some extent influenced by populations and needs and wages for staff (e.g. higher wages in London), but generally a greater fall in spending in more deprived areas between 2010/11 and 2015/16.[6]

**Appendix S2. Supplemental Methods**

Data on early childhood development for 153 English local authorities was obtained using publicly available data on child development at 2 to 2.5 years, from the Health Visitor Service Delivery Metrics, published by the Office of Health Improvement and Disparities.[7] LA boundary changes meant that two new LAs (Cumberland, Westmorland and Furness) were created in 2023/24, which we excluded. Due to small population sizes, numbers for City of London was combined with Hackney, and Isles of Scilly was combined with Cornwall. Data was reported quarterly and summarised to annual data. For a few local authorities reporting data for at least three quarters of the year, this was scaled to annual data. Each year started in April and ended in March the next year.

This dataset was published as experimental statistics from 2017/18 to 2021 and as official statistics from 2021/22.[8] Data quality and coverage cited as reasons for not being published as official statistics. There were progressive improvements in data quality and coverage despite disruptions to service delivery during 2019/20 to 2020/21. However, apart from 2017/18, data from subsequent years was comparable in quality and coverage.[9] Therefore, we only excluded 2017/18 data from analysis.

***COVID-19 time shocks***

Our within-between models enable inclusion of dummy variables for year to control for annual time shocks including due to COVID-19, enabling us to isolate the effect of within-LA changes in our predictor of interest that are not due to COVID-19 time shocks. However this approach meant that we could not capture changes in child poverty due to COVID-19 time shocks.

This assumes that COVID-19 shocks affected all local authorities to the same extent. This assumption is likely violated to the extent that COVID-19 rates varied across local authorities, and measures were not applied in the same way nationally. Most restrictions were nationally applied apart from September and October 2020, and December 2020.[9] During this period, LAs were assigned restrictions according to a tiered system and were moved into stricter restrictions as infection levels increased, until a national lockdown was applied in January 2021.

As we are also interested in effects of our predictors on the levels of the outcome between LAs, we need to control for between-LA variation in the experience of COVID-19 shocks, which may influence estimates of the levels of poverty/deprivation and child developmental outcomes for each local authority. Therefore, we used excess mortality rates to proxy the differential impact of COVID-19 lockdowns between local authorities during this period. It is a good proxy as death rates themselves are not expected to affect the outcome (death occurs after early childhood development), unless it is associated with the impact of restrictions on child development.

**Appendix S3. Within-between model**

Define i the local authority and t the birth year. Define $Dev_{it}$ the rate of children who do not achieve a good level of development, expressed in rate per 100, $x_{it}$ the vector of time-varying explanatory variables which include a full set of time dummies for t = 2018, . . . , 2022, with t = 2017 as the reference year, and $z_i$ the vector of time-constant factors of interest, which include LA ethnic composition and excess mortality during Tiered lockdown restrictions. Define the within-between model:

$$Dev_{it} = \alpha + (x_{it} - x_i)\beta_1 + x_i\beta_2 + z_i\gamma + \mu_i + \varepsilon_{it} \quad [1]$$

where $\mu_i + \varepsilon_{it}$ is the random-effect error term. The estimates of $\beta_1$ based on equation [1] are equivalent to the estimates based on standard fixed-effects models and so robust to unobserved time-constant factors.

We estimate two regressions models based on equation [1], depending on the explanatory variables included in $x_{it}$ :

- In the first model, we estimated the association between child poverty independent of children services spending (and vice versa). We include in vector $x_{it}$ the percentage of child poverty before housing costs, the total spending per child aged 0 to 5 years and the percentage of eligible population receiving an ASQ review.
- In the second model, we estimated the association of child poverty unadjusted for children services. We include in vector $x_{it}$ the percentage of child poverty before housing costs and the percentage of eligible population receiving an ASQ review.

In sensitivity analyses, we re-ran the first model using child poverty after housing costs and free school meal eligibility, in place of child poverty before housing costs.

6 Action for Children, National Children's Bureau, The Children's Society. Turning the tide. Children's Charities Coalition, 2017.

7 Office for Health Improvement and Disparities. Child development outcomes at 2 to 2 and a half years: quarterly data for 2021 to 2022. 2022; published online Aug 2. https://www.gov.uk/government/statistics/child-development-outcomes-at-2-to-2-and-a-half-years-quarterly-data-for-2021-to-2022 (accessed Nov 14, 2024).

8 Chua YW, Murray C, Munford L, Bennett D, Hargreaves D, Taylor-Robinson D. Time trends and area-level socioeconomic inequalities in early childhood development at 2 to 2.5 years old in England between 2019 and 2024. 2025; : 2025.03.14.25323968.

9 Baker C, Kirk-Wade E, Brown J, Barber S. Coronavirus: A history of English lockdown laws. 2024; published online Nov 13. https://commonslibrary.parliament.uk/research-briefings/cbp-9068/ (accessed Nov 14, 2024).
